# Establishing a framework for human dose prediction in anti-tuberculosis drug development

**DOI:** 10.64898/2026.05.26.26354063

**Authors:** Anu Patel, Angela T. Li, Belén P. Solans, Radojka M. Savic

**Affiliations:** Department of Bioengineering and Therapeutic Sciences, University of California San Francisco, San Francisco, CA, USA; UCSF-UC Berkeley Joint PhD Program in Bioengineering, University of California San Francisco, San Francisco, CA, USA

**Keywords:** Tuberculosis, Translational Science, Dose-Response Relationship

## Abstract

**Rationale:** Efficacious dose selection for anti-tuberculosis drugs has traditionally relied on achieving plasma exposures above the minimum inhibitory concentration, but this approach has not consistently aligned with clinical outcomes.

**Objectives:** We sought to identify early pharmacokinetic-pharmacodynamic targets most predictive of clinical efficacious dose.

**Methods:** We conducted a back-translational, pharmacokinetic-pharmacodynamic simulation-based analysis of 15 anti-tuberculosis drugs. Using pharmacokinetic data from multiple biological matrices and a range of pharmacodynamic metrics, we established candidate exposure-response targets for attainment. We systematically evaluated the predictive accuracy of each target pair against established clinical doses to formulate a decision-making framework linking key drug properties to the most predictive targets.

**Measurements and Main Results:** Depending on the target used, projected clinical doses varied widely—both within and across compounds—highlighting the importance of target selection for dose projection and go/no-go decisions. In general, targeting cellular lesion-level drug exposures relative to *in vivo* preclinical potency provided an effective approach for early dose selection. However, for highly penetrating drugs, targeting site-of-action therapeutic exposures in the caseum was more predictive of clinical dose. Based on these findings, we developed a preliminary dose prediction tool that enables drug developers to estimate clinically relevant dose ranges of compounds using *in vitro* and early *in vivo* data.

**Conclusions:** This work establishes and validates a simple, evidence-based framework to standardize early translational decision-making on dose selection of anti-tuberculosis candidates in development.

## Introduction

Approximately 40% of all candidates in drug development are lost due to inappropriate metabolism and pharmacokinetic (PK) profiles,^1^ and the main cause of failure in clinical trials is poor efficacy.^2^ Exposure-response relationships help inform the clinical dose of a compound in development. Being able to effectively project clinical PK and efficacy from data produced in early stages of development is imperative to determine efficacious doses, recognize potential issues early, and make better-informed decisions to prioritize the most promising candidates for progression.

Identification of efficacious dose is particularly important in the field of tuberculosis (TB), given the high global burden of the disease and lengthy process of regimen development. Approximately 1.3 million deaths are caused by TB annually despite the availability of a 6-month standard-of-care regimen,^3^ motivating the focus of current efforts on shortening treatment durations to increase adherence and improve outcomes.^4–7^ However, anti-TB regimen development is long, in part due to the Phase 3 endpoint of treatment failure or relapse, which can be measured up to 12-18 months post-treatment.^8^ Notably, if a suboptimal dose of a novel compound is used in a combination regimen tested in a Phase 3 clinical trial, this could decrease the likelihood of trial success, leading to inefficiencies in resources and time.

Dose selection may be driven by a goal such as maintaining drug levels above an efficacy target for a dosing period (i.e., attainment of a pharmacokinetic-pharmacodynamic (PK-PD) target).^9,10^ Although an *in vitro* cutoff such as the minimum inhibitory concentration (MIC) is widely considered to be an acceptable target for dose optimization and improved treatment response,^11–15^ integrated PK and microbiology-based PD measures tested in late-stage clinical trials to confirm relationships are seldom performed. Indeed, both the PK and PD components of this target are limited in recapitulating the complex nature of the disease and pathogen. Recent work has shown site-of-action PK as a valuable facet for understanding drug efficacy in hard-to-treat patients with cavitary disease.^16^ Furthermore, the MIC is determined in a biologically irrelevant matrix, selects for actively replicating bacteria, and indicates growth inhibition rather than killing, which is needed for relapse-free cure.^17^

Currently, there are 15 anti-TB drugs that are either approved for the disease or are in clinical development: bedaquiline (BDQ), BTZ-043, delamanid (DLM), ethambutol (EMB), isoniazid (INH), linezolid (LZD), moxifloxacin (MXF), pretomanid (PMD), pyrazinamide (PZA), rifampicin (RIF), rifapentine (RPT), sutezolid (SZD), TBAJ-587, sorfequiline (TBAJ-876), and TBI-223. Using data from these drugs, we performed a back-translational modeling analysis, aiming to create a framework for human dose prediction through the identification of appropriate PK-PD targets based on *in vitro* and early *in vivo* measures produced in drug development. We assessed the accuracy of the predicted doses from each PK-PD target by benchmarking them against their known clinically efficacious doses. Finally, we developed a dose projection tool based on our findings to apply to new compounds in development.

## Methods

### Pharmacokinetic data collection

Clinical plasma concentrations were quantified from population PK models developed in-house or sourced from literature **(Table S1)**. Clinical site-of-disease concentrations **(Table S2)** were quantified from a translational site-of-disease PK modeling platform previously developed by our group.^16^ Unbound concentrations were utilized when comparing against unbound metrics (i.e., plasma PK over MIC); otherwise, total concentrations were used.

### Pharmacodynamic data collection

*In vitro* PD metrics assessed were the minimum inhibitory concentration (MIC), the concentration at which 90% of bacterial growth is inhibited in the macrophage (macIC_90_), the concentration that kills 50 or 90% of non-replicating persisters in *ex vivo* caseum (casMBC_50/90_), and the *in vivo* effective concentration (EC_50/90_) **(Table S4)**. MIC values were assessed in standard media (7H9 broth, or 7H10, 7H11, or 7H12 medium), with or without enrichment. MacIC_90_ values were measured in human monocyte-derived macrophages, and casMBC values were quantified in *ex vivo* caseum from New Zealand White rabbits. *In vivo* EC_50/90_ values were quantified for each drug using a BALB/c mouse-to-human translational PK-PD modeling platform previously built in our group.^18^ For clinical translation, EC_50/90_ values were corrected by interspecies plasma protein binding differences. For drugs in which plasma protein binding exceeded 99% in both mice and humans, the interspecies binding ratio was assumed to be 1. Unbound fractions in plasma for each drug in each species are detailed in **Tables S1** and **S7**.

### Pharmacokinetic-pharmacodynamic coverage simulations

Plasma exposures from a dose range of 0.1 to 10000 mg were simulated for each drug using the clinical population PK models. Site-of-disease exposures were simulated using estimated partitioning and delay rates from plasma to each site-of-disease **(Table S3)**.

Coverage was defined as the time over which PK is above the relevant PD target for each PK matrix. Each matrix was compared to the most pertinent biological target for a given microenvironment. Specifically, drug exposures in uninvolved lung and cellular lesions were compared to macIC_90_, as these house intracellular bacteria in macrophages and other immune cells. Caseum exposures were compared to casMBC_90_, and those in plasma to MIC, which is the standard clinical comparison for efficacy. Cellular lesion exposures were compared to *in vivo* EC_50_, as the cellular lesions represent a site-of-disease in BALB/c mouse models, which do not form necrotizing lesions. Coverage was assessed at a 24-hour interval at steady-state. For each drug, the predicted effective dose range was defined as doses producing 50 or 90% of coverage (i.e., time above target) during a 24-hour interval at steady-state. The predicted doses from each PK-PD target were compared to the highest current effective dose of each drug, or the median dose if there is a range, as determined through clinical trial results and/or modeling analyses **(Table S5)**. Accurate projection was defined as being within two-fold of the current efficacious dose.

### Software and statistical analysis

PK simulations were performed in NONMEM (v7.5). The human dose prediction tool was built using R package *shiny* and utilized *mrgsolve* for simulations. PK simulations were based on typical parameter estimates and did not account for residual error or parameter uncertainty.

Further methodological details are available in the Online Supplement.

## Results

### Efficacious dose projection using pharmacokinetic-pharmacodynamic coverage

To project the clinical dose of anti-TB drugs, several PK and PD measures can be used, which increase in relevance and complexity throughout development (**Figure 1**). For this work, we specifically focused on assessing data that is commonly collected for developmental compounds; however, to reduce uncertainty in early PK projection, clinical PK was used. Overall, predicted doses of an individual compound varied widely depending on the PK-PD target used, highlighting the inconsistency in decision-making that may occur depending on the selected target. For instance, for some drug classes like the diarylquinolines and rifamycins, only one PK-PD target was predictive of efficacious dose. However, for other classes such as the oxazolidinones, multiple targets performed similarly well. Furthermore, dose predictions differed across compounds, showcasing the high level of variability between anti-TB drugs. For example, dose predictions using unbound plasma PK over MIC ranged from a 12-fold underprediction to a 54-fold overprediction, depending on the drug (**Figure 2**).

**Figure 1.**
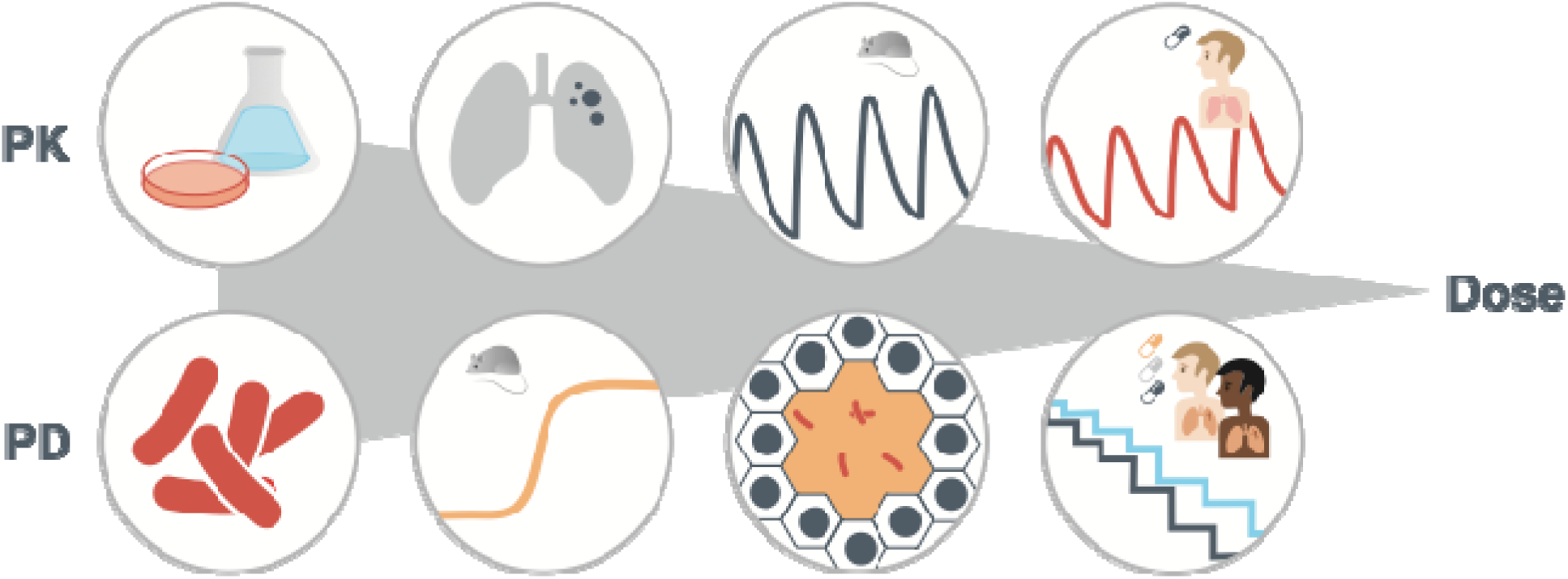
Overview of human dose prediction for anti-tuberculosis compounds. Several pharmacokinetic (PK) and pharmacodynamic (PD) data are produced for a new compound throughout different stages of development. Initially, hepatocyte or microsomal stability assays are used to quantify *in vitro* PK, and potency assays are used to determine *in vitro* PD. Site-of-action PK-PD is later determined through evaluating drug distribution and efficacy in tissues. Preclinically, plasma PK can be collected for various species, and a PK-PD relationship can be determined through modeling dose-ranging data in mouse. Finally, exposure-response is determined in late-stage clinical trials.

**Figure 2.**
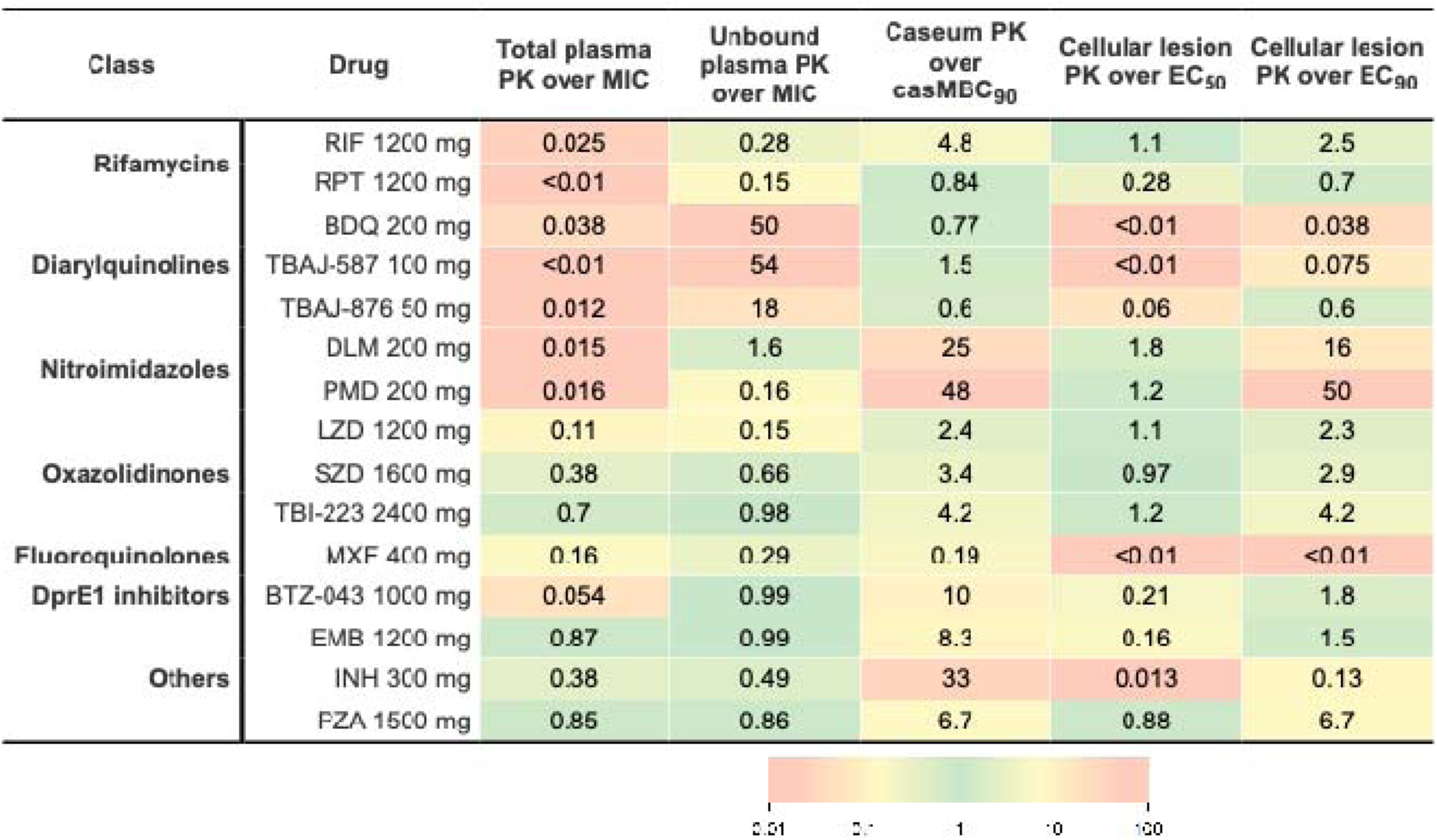
Ratio of predicted ED_50_ to clinical efficacious dose across all PK-PD targets. Ratios were log_10_-transformed and mapped to a symmetrical green-to-red color scale. Values close to one are shown in green, indicating minimal deviation between predictions and observations. Large deviations in either direction are shaded in red.

### Recommendations for dose projection methods of new drug candidates

Variation in PK and PD data availability across preclinical stages was used to inform the development of the dosing framework. In the lead optimization stage, usually only preclinical plasma PK and *in vitro* PD (i.e., MIC) are available. In this stage, total plasma PK over MIC was generally poorly predictive of clinical dose, with 12 of 15 drugs being underpredicted by 2.5- to over 100-fold when comparing projected ED_50_ to the clinical dose. Correcting exposures with protein binding by using unbound plasma PK over MIC led to accurate predictions for BTZ-043 and SZD but still underpredicted the dose of 6 drugs. On the other hand, this metric overpredicted doses of highly bound compounds (i.e., those with plasma protein binding >99.9%, all belonging to the diarylquinoline class, **Table S1**) by 18- to 54-fold.

At the selection phase, lesion-level exposure and efficacy data typically become available. For highly penetrating drugs (i.e., those with a ratio of predicted average steady-stage concentration (Css,avg) in clinical caseum to casMBC_90_ of approximately one or higher) **(Table S6)**, 4 of 5 were predicted accurately using caseum PK over casMBC_90_, with MXF being underpredicted. At the preclinical candidate stage, dose-ranging murine PK-PD studies are conducted. Based on mouse PK simulations using human-equivalent doses **(Table S7** and **S8)**, for highly potent drugs (i.e., those with a mouse plasma Css,avg at least three-fold higher than the estimated *in vivo* EC_50_) **(Table S9)**, cellular lesion PK over EC_50_ underpredicted dose (by over 3.5-fold) for all 8 compounds. For the remaining drugs for which mouse plasma Css,avg was comparable to human exposures, cellular lesion PK over EC_50_ accurately predicted the dose of all 7 compounds (**Figure 2**). A similar trend was seen when comparing projected ED_90_ against the clinical dose **(Table S10; Figure S1)**.

As preclinical development advances, dose projection becomes more precise, given that more data is available to make more informative decisions. Thus, at the latest stage, when all preclinical data is available, cellular lesion PK over *in vivo* EC_50_ is an appropriate PK-PD target for dose-related decision-making. Exceptions include drugs that are either highly potent (i.e., those having high average plasma exposures relative to *in vivo* half-maximal potency) and/or highly lesion-penetrating (i.e., those having higher average caseum exposures relative to potency against non-replicating persisters). Out of these exceptions, those that are highly penetrating (BDQ, MXF, RPT, TBAJ-587, and TBAJ-876) are generally well-predicted using caseum PK over casMBC_90_ as a target. For those that are highly potent but less or non-penetrating (BTZ-043, EMB, and INH), using cellular lesion PK over a stricter target, the EC_90_, is more appropriate. When applying this framework, we accurately predicted the efficacious doses of 13 of 15 drugs and underpredicted dose for two drugs (INH and MXF) (**Figure 3**).

**Figure 3.**
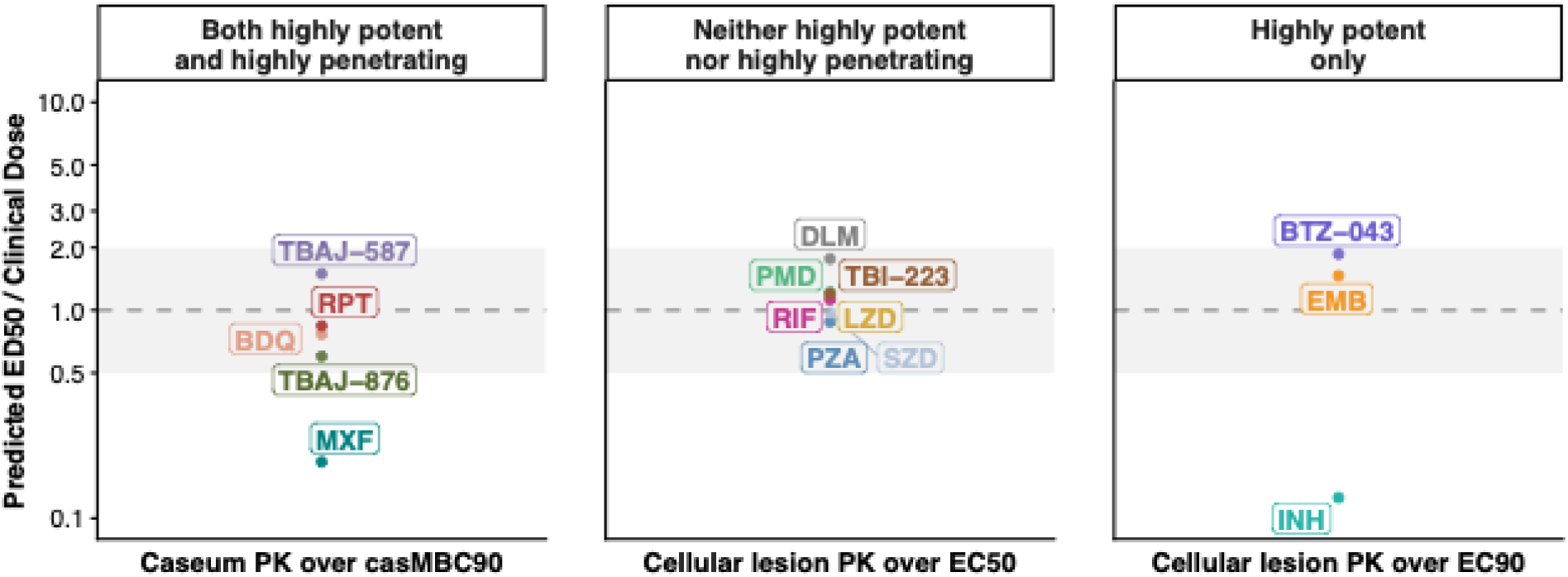
Drug categorization based on findings from PK-PD target comparison. Horizontal grey dashed line is the line of unity, representing exact prediction of clinical dose. Light grey shaded area represents a two-fold deviation from the line of unity.

### Clinical efficacious dose prediction framework and tool

Based on the observed findings, a decision-making framework was created to guide drug developers in making more informative decisions regarding compound selection and prioritization at various stages of preclinical development (**Figure 4**). This schema provides a dose projection pathway that utilizes simple rules-of-thumb based on key data collected in respective phases of early development. A simulation-based web tool was developed to understand dosing predictions using various PK-PD targets (**Figure 5**). Using this, a developer can input the data available for a candidate compound and project dose. As clinical PK would be unavailable in preclinical development, *in vitro-in vivo* extrapolation and allometric scaling, methods commonly used to scale *in vitro* and preclinical PK to human estimates, were incorporated in the dose prediction tool. User-preferred scaling methods can be chosen to project clinical PK and calculate coverage over *in vitro* or *in vivo* PD targets.

**Figure 4.**
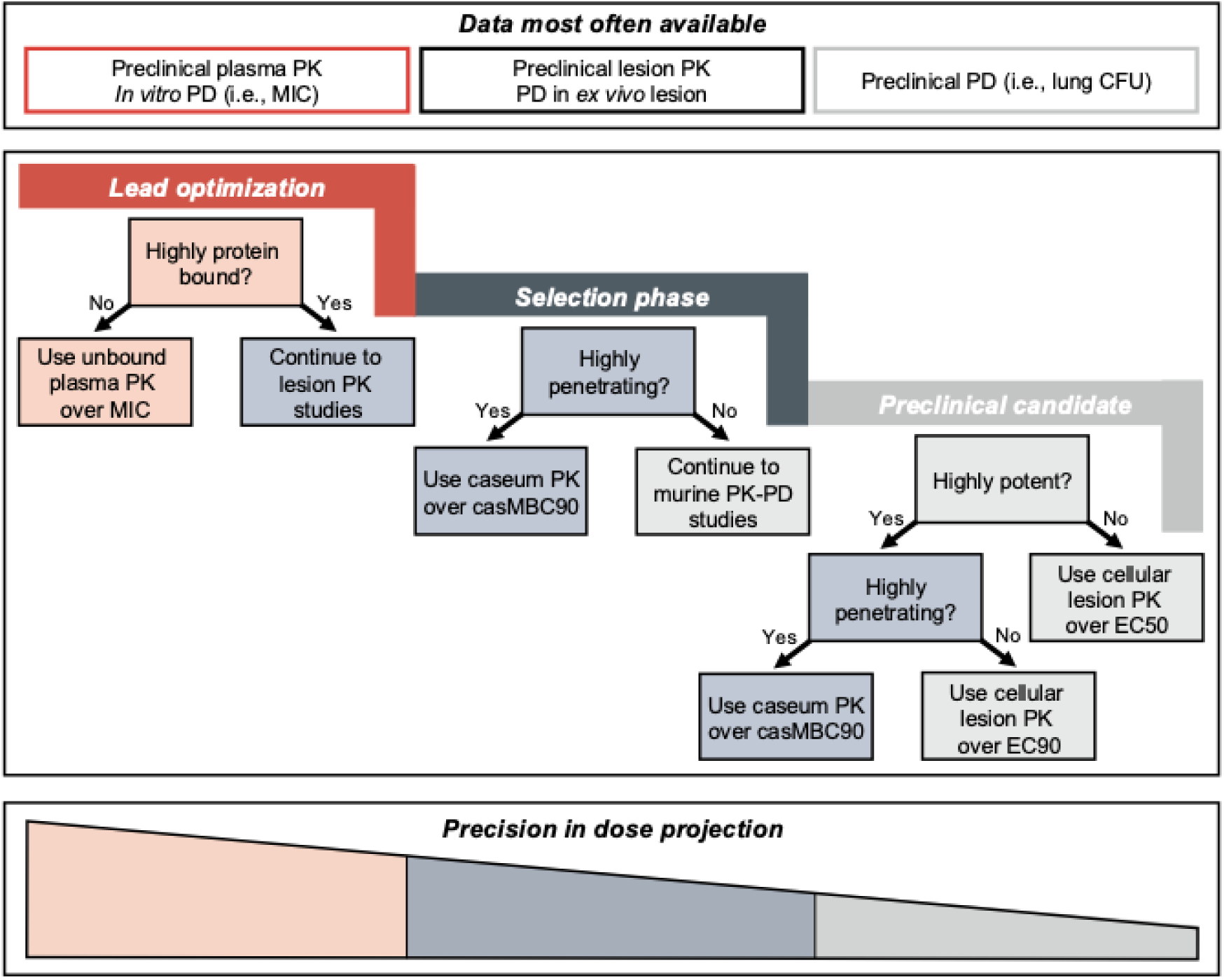
Decision-making framework for dose projection of anti-TB drugs. At different stages of development, specific types of data are generally outputted for a particular compound (top). This data can be incorporated to create dose projections across each stage (middle). As more data becomes available, precision in the projected dose based on drug characteristics increases, until the preclinical candidate stage is reached, where the most informative decision can be made (bottom). Highly protein bound is defined by having a plasma protein binding of >99.9%. Highly penetrating is defined by having a caseum Css,avg to casMBC_90_ ratio of ≥1. Highly potent is defined by having a mouse plasma Css,avg to *in vivo* EC_50_ ratio of >3. *Abbreviations:* Css,avg: average steady-state concentration.

**Figure 5.**
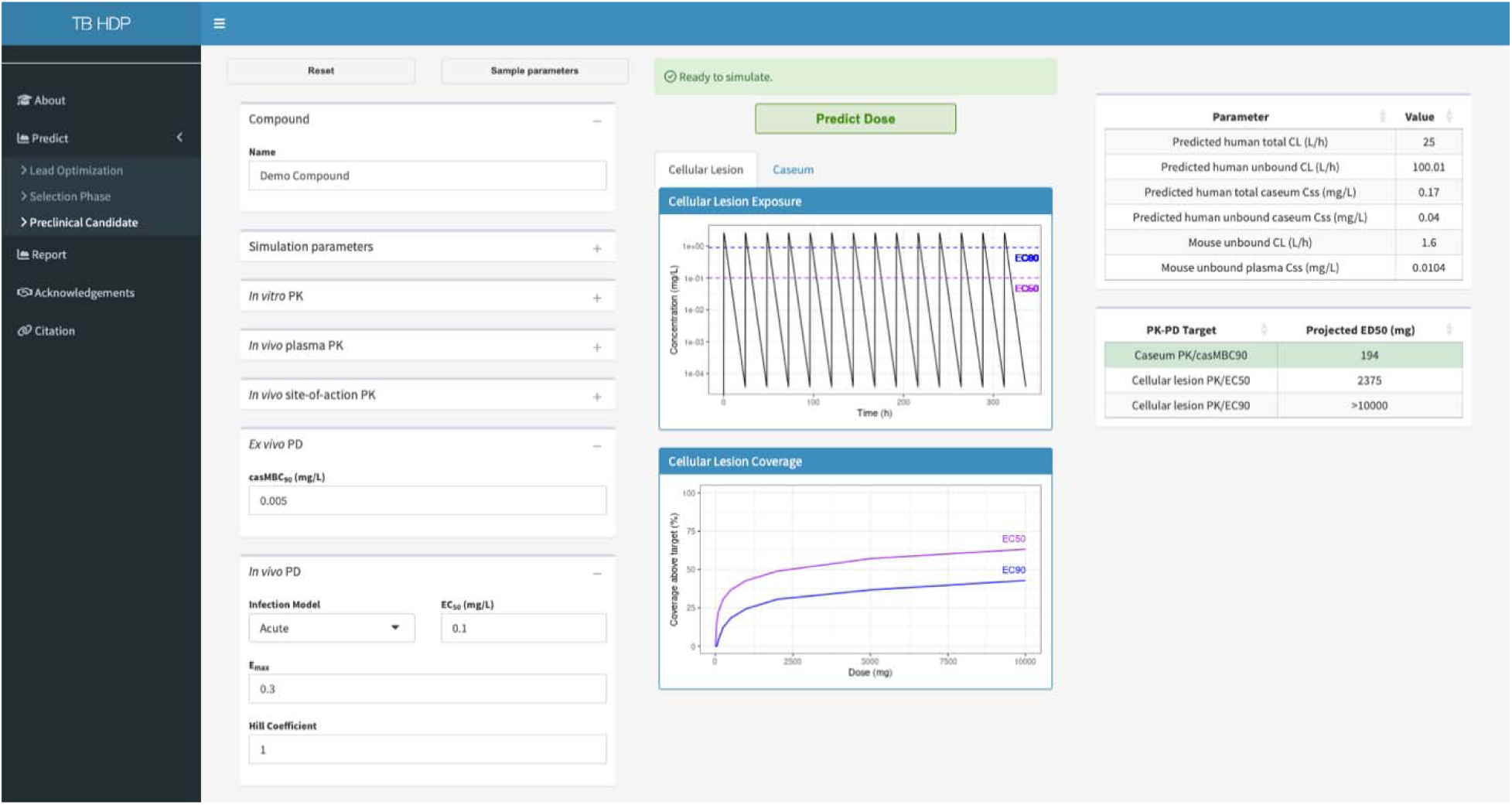
Interface of dose prediction tool for anti-TB compounds in development. Users can input specific *in vitro* and *in vivo* data for anti-TB compounds to project clinical PK and efficacious dose. This tool can be accessed at: http://angelatli.shinyapps.io/Human-Dose-Prediction/.

## Discussion

Accurate clinical dose projection, defined by PK, efficacy, and safety properties, is vital for a drug candidate’s success. However, issues related to appropriate dose determination can arise even after approval. A review of 218 approved compounds by the U.S. Food and Drug Administration (FDA) between 2018 to 2022 showed that 28 (13%) had modifications to the proposed dose or needed additional post-marketing assessments. Specifically, it revealed that, (1) observing a dose/exposure-response relationship for safety, but not efficacy, was associated with an increased probability of downward dose modifications, (2) the use of PD biomarkers for dose setting was associated with decreased probabilities of downward change, and (3) drugs indicated for infectious diseases underwent more upward modifications than those in other therapeutic classes.^19^

In TB, similar observations were shown with rifampicin, a critical drug in the standard treatment of drug-susceptible TB dosed at 10 mg/kg (typically 600 mg) in adults.^20^ This dose was initially chosen in the 1980s in part because early PK studies established that a once-daily administration of 600 mg yielded serum concentrations well above the minimum inhibitory concentration (MIC) of the causative pathogen, *M. tuberculosis*.^21^ Only recently have PK-PD modeling and clinical studies testing higher doses of the drug shown that the historically-recommended dose indeed may not produce maximal therapeutic benefit.^22,23^ Later, the dose of rifapentine was initially chosen to replicate that of rifampicin,^24^ but it was only after conduction of a Phase 2 dose-ranging study that higher doses of rifapentine were shown to have higher bactericidal activity.^25^ These are two examples of how the identification of optimal doses/exposures is key to treatment success.

The FDA has provided a Guidance for Industry on drug development for pulmonary TB, stating that, when selecting a dosing regimen to be evaluated in Phase 3 clinical trials, sponsors should consider target PK-PD parameters (e.g., area under the curve/MIC, maximum concentration/MIC, time above the MIC) based on *in vitro* models and animal models of TB, results from early clinical trials, and results from exposure-response evaluations.^26^ This broad statement highlights the need for a standardized dose projection framework. Furthermore, while more complex methods can be utilized, PK-PD target attainment is an accessible and simple method for drug developers that allows for preliminary decision-making using experimental data produced early in development.

Dose optimization has also been emphasized in other fields, such as oncology, with the launch of Project Optimus by the FDA in 2021. This initiative emphasizes that dose selection should prioritize optimizing efficacy, rather than solely relying on the traditional maximum tolerated dose approach. Key considerations include determining whether there are any PD biomarkers that may help inform dose optimization, integrating modeling and simulation with emerging clinical data to support dose optimization, planning dose-optimization strategy early, and collecting exposure data from multiple dosages to gain better understanding of exposure-efficacy relationships, all of which were applied in this current work.^27^

Here, through a back-translational analysis, we projected the doses of 15 anti-TB drugs using several different PK-PD targets. We benchmarked these projections against the clinical efficacious dose of each drug as identified through clinical studies and/or modeling analyses. Finally, we systematically assessed dose projections across targets to formulate a decision-making framework for identifying clinical dose in early development. Overall, we showed that, contrary to its prevalence in the field, plasma PK over MIC is poorly predictive of clinical dose and is not a generalizable metric for early developmental decision-making. Even when correcting for protein binding, this metric significantly penalized highly protein-bound drugs. Such systematic bias risks the premature de-prioritization of highly efficacious drugs, such as the diarylquinoline class. In contrast, more accurate dose projections can be achieved by integrating individual drug properties, such as plasma protein binding, extent of lesion penetration, and potency, with drug concentrations measured in biologically-relevant matrices that reflect the site of disease and optimal drug effect.

Specific categorizations were developed to build this decision-making framework with regards to identifying dose. It is important to note that the spectrum of drug activity is wide—although a drug may not be considered “highly potent” in this context, this does not denote that the drug does not have appropriate potency to achieve bactericidal effect. Similarly, a drug not considered “highly penetrating” does not indicate that it is not effective in the lesions. Nevertheless, to add to the confidence in this classification, categories aligned well with drug class, as drugs in the same class exhibited similar trends with regards to the predictability of certain PK-PD targets. From this categorization, we accurately predicted doses (within two-fold) for 13 of 15 drugs, but INH and MXF were significantly underpredicted. Notably, INH is a historical anti-TB drug, and MXF at its dose was repurposed from other infectious disease indications. In the simulations, we utilized available clinical PK to reduce uncertainties surrounding early PK projection. Several PK scaling methods are currently being utilized in the field, with varied consensus on their accuracy and relevance. Each method has been shown to have use cases where projection is more accurate, as well as cases where projections are limited, depending on a drug’s PK properties.^28–30^ Thus, we incorporated scaling in our prediction tool to allow the user to utilize any approach to quantify clinical PK when this is not yet available.

This work is subject to certain limitations. First, the focus of this work is on identifying efficacious dose and did not consider safety, which is an important aspect of dose determination. However, anti-TB drugs historically have not undergone dose modifications due to safety reasons, with an exception being linezolid, which was tested at lower doses and durations in subsequent clinical trials due to toxicity concerns.^31^ Second, PK simulations were performed across a wide dose range (up to 10000 mg). If linear PK was identified for a particular drug based on the dose range for which the PK model was built, this assumption may not hold for higher doses, resulting in potential overprediction of exposures, and therefore underprediction in dose. Third, our goal was to compare biologically-relevant PD targets against PK matrices, ensuring appropriate alignment of measurements (i.e., total-to-total or unbound-to-unbound). In this work, we assumed MICs to reflect unbound concentrations as they were measured in standard, and not protein-supplemented, media; however, the presence of some protein or components leading to non-specific binding can still impact free drug concentrations and influence projected doses. Fourth, anti-TB compounds are given as part of a combination treatment comprising three to four drugs. This work assumed that the effective dose of a drug does not vary when given in different drug combinations (i.e., it did not consider synergistic or antagonistic effects) which follows the current process by which anti-TB drugs are generally developed. Finally, the decision-making framework was built using retrospective data on a limited number of drugs found to be efficacious.

We attempted to enhance our dataset with drugs currently in development, with the recognition that these doses are not final. Some drugs may have achieved their doses due to the historical thinking that plasma PK over MIC is a reliable method—including the historical drugs EMB and PZA, both of which were well-predicted by using this metric. Thus, some conclusions could be confounded by the fact that higher doses could be more effective, but these were never tested. Data for drugs that were unsuccessful in reaching clinical stages was not available; including such data would have further strengthened this work. To support and improve these findings, future work will focus on incorporating prospective dose predictions of new drug candidates into this framework.

## Conclusion

In totality, this work creates a simple guidance—based on data commonly collected early in development—on performing clinical efficacious dose projection of anti-TB drug candidates. We showcase the value of using biologically- and clinically-relevant PK-PD metrics to optimize efficacy in dose projections throughout preclinical development, with the aim to standardize and inform dose-related translational decision-making and increase the chances of regimen success.

## Data Availability

All data utilized in this work is detailed in the Online Supplement. The code used to build the dose projection tool is found at https://github.com/angelatxli/TB-HDP.

## Acknowledgments

We would like to thank Mike Morimoto and Griffith Gao for their technical support in the implementation of the human dose prediction tool.

