## Supplemental for "Establishing a framework for human dose prediction in anti-tuberculosis drug development"

**ONLINE SUPPLEMENT**

**SUPPLEMENTARY METHODS**

**Pharmacokinetic data collection**

Clinical plasma concentrations were quantified from population PK models either built in-house (Savic laboratory at UCSF, unpublished) or taken from literature, described in **Table S1**. Clinical site-of-disease concentrations **(Table S2)** were quantified from a translational site-of-disease PK modeling platform, which was previously developed in our group and described in detail in [^S1^]. To summarize, this platform was built from plasma and tissue PK data collected in uninfected and infected New Zealand White (NZW) rabbits for 24 different drugs and drug candidates. Tissue concentrations were taken from rabbits at three main sites of disease: uninvolved/non-diseased lung tissue, cellular lesions, and caseum. All PK samples (collected as either homogenates or through laser capture microdissection) were analyzed using liquid chromatography tandem mass spectrometry. A modeling structure was built in which plasma PK was linked to tissue concentrations, and the rate ($k_{pt}$) and extent ($PC$) of drug partitioning was estimated from plasma to each tissue type. After determining the best-fitting plasma model, individual and population plasma PK parameters were fixed, and population tissue PK parameters were estimated. The change in drug concentrations over time in a given tissue type is described in **Equation S1**.

$\frac{{dC}_{tissue}}{dt}=k_{pt}\times\left( {PC}_{tissue}\times C_{plasma}- C_{tissue} \right)$	**(Eq. S1)**

$k_{pt}$, rate of delay between plasma and tissue type; ${PC}_{tissue}$, partition coefficient between tissue type and plasma; $C_{plasma},$ plasma drug concentration; $C_{tissue}$, drug concentration within a tissue type.

Human site-of-disease PK models were similarly built from clinical tissue data (NCT00816426) for seven overlapping drugs in rabbits and humans (clofazimine (CFZ), INH, kanamycin (KAN), LZD, MXF, PZA, and RIF). Partitioning in humans and rabbits for these seven drugs was compared, and an interspecies partitioning relationship was quantified **(Equation S2).** For the remaining 17 drugs not studied in humans, rabbit partition coefficients for each tissue type were calibrated using this interspecies partitioning relationship, in order to reflect human partitioning. Calibrated or human partition coefficients and delay rates can be found in **Table S3**.

${PC}_{H}= {-0.18+0.58\cdot PC}_{R}$	**(Eq. S2)**

${PC}_{H}$, partition coefficient in human; ${PC}_{R},$ partition coefficient in rabbit. PC values are in log_10_.

Plasma protein binding values were either identified in literature, provided by asset owners, or provided by collaborators **(Table S1)**. Unbound concentrations were utilized when comparing against unbound metrics (i.e., plasma PK over MIC); otherwise, total concentrations were used.

**Pharmacodynamic data collection**

Potency metrics and references for each drug can be found in **Table S4**. *In vitro* PD metrics assessed were the MIC, macIC_90_, and casMBC_50/90_; all values have been previously published and are described in [^S1^]. MICs were assessed in standard media (7H9 broth, or 7H10, 7H11, or 7H12 medium), with or without enrichment. MacIC_90_ values were quantified in human monocyte-derived macrophages, and casMBC values were quantified in *ex vivo* caseum from NZW rabbits.

The *in vivo* EC_50/90_ was also assessed, which was quantified for each drug using a translational PK-PD modeling platform previously built in our group and described in detail in [^S2^]. Briefly, this platform, built from plasma PK and longitudinal colony-forming unit (CFU) data in BALB/c mice, describes collective mycobacterial dynamics as a result of natural growth, mouse immune response, and drug effect.

To build this platform, first, mouse PK models were developed to quantify concentrations to drive drug effect **(Table S7)**. Next, to estimate the drug effect independent of natural bacterial dynamics, a disease progression model was applied to quantify mouse immune response to infection with *M. tuberculosis* in BALB/c mice.^S3^ In this model, the effect of the mouse immune system on bacterial growth was found to depend both on the infection duration, as adaptive immunity requires time to reach full effect, and the initial inoculum size, where a larger infection allows the bacterial number to more quickly approach a tipping point at which the adaptive response is activated. The relationships between time and initial bacterial burden on bacterial growth are shown in **Equation S3**. To account for drug effect, linear, Emax, and sigmoidal Emax relationships were tested directly on the bacterial compartment **(Equation S4)**. A delayed Emax relationship **(Equation S5)** was also tested to describe the delay in concentrations from plasma to site of disease. Mouse population PK parameters were fixed to estimate effect.

$\frac{dB}{dt}=K_{g}\times B\times\left( 1-\frac{K_{B} \times B^{\gamma_{B}}}{B_{50}^{\gamma_{B}}+ B^{\gamma_{B}}} \right)\times\left( 1-\frac{{K_{T} \times t}^{\gamma_{T}}}{{T_{50}}^{\gamma_{T}} {+ t}^{\gamma_{T}}} \right) -K_{d}\times B$	**(Eq. S3)**

$\frac{dB}{dt}=K_{g}\times B\times\left( 1-\frac{K_{B} \times B^{\gamma_{B}}}{B_{50}^{\gamma_{B}} + B^{\gamma_{B}}} \right)\times\left( 1-\frac{{K_{T} \times t}^{\gamma_{T}}}{{T_{50}}^{\gamma_{T}}{+ t}^{\gamma_{T}}} \right) -K_{d}\times B-{EFF}_{drug}\times B$	**(Eq. S4)**

$\frac{dC_{delay}}{dt}=k_{delay}\times\left( C_{plasma}-C_{delay} \right)$	**(Eq. S5)**

$B$, bacterial colony-forming units (CFU); $t$, time since inoculation; $K_{g}$, bacterial growth rate (1/d); $K_{d}$, bacterial death rate (1/d); $K_{B}$, maximal inhibitory CFU-dependent adaptive immune effect, $B_{50}$, CFU counts to reach half of $K_{B}$; $\gamma_{B}$, steepness of CFU-dependent adaptive immune effect curve; $K_{T}$, maximal inhibitory time-dependent adaptive immune effect; $T_{50}$, time to reach half of $K_{T}$; $\gamma_{T}$, steepness of time-dependent adaptive immune effect curve; ${EFF}_{drug}$, drug effect (1/d); $k_{delay}$, rate of delay from plasma concentration associated with drug effect (1/d); $C_{plasma}$, plasma drug concentration (mg/L); $C_{delay}$, delayed concentration driving drug effect (mg/L); $E_{max}$, maximum drug effect, quantified as a rate constant of decline in CFU (1/d); ${EC}_{50}$, delayed concentration resulting in half $E_{max}$ (mg/L); $\gamma$, steepness of the relationship between delayed plasma concentrations and drug effect.

For clinical translation, EC_50/90_ values were corrected by interspecies plasma protein binding differences, with the assumption that at free drug concentrations in plasma, the PK-PD relationship is comparable between mice and humans. For drugs in which plasma protein binding was greater than 99% for both mouse and human, the interspecies binding ratio was considered to be 1, in order to minimize overcorrection for experimental variability. Unbound fractions in plasma for each drug in each species can be found in **Tables S1** and **S7**.

**Pharmacokinetic-pharmacodynamic coverage simulations**

Plasma exposures from a dose range of 0.1 to 10000 mg were simulated for each drug using the clinical population PK models. Site-of-disease exposures were simulated using estimated partitioning and delay rates from plasma to each site-of-disease. For drugs for which human tissue data was available, human estimates were used, otherwise, calibrated partition coefficients calculated from the interspecies partitioning relationship **(Equation S2)** were used. When only rabbit estimates were available, delay rates were assumed to be the same in humans **(Table S3)**. Coverage was defined as the time over which PK is above the relevant PD target for each PK matrix. Each matrix was compared to the most pertinent biological target for a given microenvironment. Namely, drug exposures in uninvolved lung and cellular lesion were compared to macIC_90_, as these house intracellular bacteria in macrophages and other immune cells, with a caveat being that the drug concentrations reflect both intracellular and extracellular exposures in these tissue types. Caseum exposures were compared to the casMBC_90_, and those in plasma to MIC, which is the standard clinical comparison for efficacy. Cellular lesion exposures were compared to *in vivo* EC_50_, as the cellular lesions represent a site-of-disease in BALB/c mouse models, which do not form necrotizing lesions. Plasma PK was also adjusted for protein binding differences when compared to MIC. Coverage was assessed at a 24-hour interval at steady-state (i.e., 4 months after daily dosing).

**SUPPLEMENTARY FIGURES AND TABLES**

**Table S1. Clinical PK data and models used for simulations**

| **Drug** | **Doses used in model building** | **Simulated model structure** | **Model reference** | **fu** |
| --- | --- | --- | --- | --- |
| BDQ | 400 mg daily (x 2 weeks), followed by 200 mg thrice weekly | 3 compartment disposition; transit model absorption; first-order elimination; allometric scaling by WT | ^S4^ | 0.000053 |
| BTZ-043 | 250, 500, 750, 1000, 1250, 1500, 1750 mg daily | 2 compartment disposition; transit model absorption; first-order elimination *(parameters for fasted state)* | ^S5^ | 0.055 |
| DLM | 100 mg twice daily | 1 compartment disposition; transit model absorption; first-order elimination | ^S6^ | 0.005^S7^ |
| EMB | 800, 1000, 1200, 1500 mg daily | 2 compartment disposition; transit model absorption; first-order elimination | ^S8^ | 0.879^S9^ |
| INH | 100, 225, 240, 300, 400 mg five days/week; 200, 300, 450 mg daily | 2 compartment disposition; first-order absorption with lag time; first-order elimination; allometric scaling by WT *(parameters for intermediate / fast eliminators)* | ^S10^ | 0.862^S9^ |
| LZD | 600 mg twice daily, 1200 mg daily | 2 compartment disposition; first-order absorption; Michaelis-Menten elimination | ^S11^ | 0.69^S12^ |
| MXF | 400 mg daily | 2 compartment disposition; transit model absorption; first-order elimination | In-house | 0.55^S13^ |
| PMD | 50, 200, 250, 400, 500, 750, 1000, 1250, 1500 mg once; 50, 100, 150, 200, 600, 800, 1000, 1200 mg daily | 1 compartment disposition; transit model absorption; first-order elimination; allometric scaling by WT; decreasing F with dose ≥200 mg | ^S14^ | 0.14^S15^ |
| PZA | 1000, 1500, 2000 mg daily | 1 compartment disposition; transit model absorption; first-order elimination; decreasing F with dose >1000 mg | ^S16^ | 0.99^S9^ |
| RIF | 10, 20, 25, 30, 35, 40 mg/kg daily | 1 compartment disposition; transit model absorption; autoinduction via an enzyme turnover model; increasing F with dose >450 mg; allometric scaling with FFM | ^S17^ | 0.118^S9^ |
| RPT | 1200 mg daily | 1 compartment disposition; transit model absorption; first-order elimination | In-house | 0.003 |
| SZD | 300, 600, 1200, 1800 mg once | 2 compartment disposition; first-order absorption; decreased F with 1800 mg dose; Michaelis-Menten elimination | In-house | 0.469 |
| TBAJ-587 | 25, 50, 100, 200, 400, 800 mg once; 25, 75, 200 mg daily | 3 compartment disposition; transit model absorption; first-order elimination | ^S18^ | 0.00034 |
| TBAJ-876 | 10, 25, 50, 100, 200, 400, 800 | 3 compartment disposition; transit model absorption; first-order elimination | ^S19^ | 0.00043 |
| TBI-223 | 50, 100, 300, 600, 1200, 1800, 2000, 2600 mg once | 1 compartment disposition; transit model absorption; Michaelis-Menten elimination *(parameters for sustained-release formulation)* | ^S20^ | 0.634^S20^ |

For models with allometric scaling, a typical WT of 60 kg, FFM of 45 kg,^S21^ and FAT of 15 kg were used.

*Abbreviations:* F: bioavailability, FAT: fat mass, FFM: fat-free mass, fu: fraction unbound in plasma, WT: body weight.

**Table S2. Clinical PK measures**

| **Drug** | **Dose (mg)** | **Cmin (mg/L)** | | | | | **Css,avg (mg/L)** | | | | | **Cmax (mg/L)** | | | |
| --- | --- | --- | --- | --- | --- | --- | --- | --- | --- | --- | --- | --- | --- | --- | --- |
|  |  | **Plasma** | **Uninvolved lung** | **Cellular lesion** | **Caseum** | **Plasma** | | **Uninvolved lung** | **Cellular lesion** | **Caseum** | **Plasma** | | **Uninvolved lung** | **Cellular lesion** | **Caseum** |
| BDQ | 200 | 1.54 | 17.4 | 14.5 | 2.58 | 1.90 | | 21.5 | 16.1 | 3.18 | 2.33 | | 26.2 | 17.3 | 3.89 |
| BTZ-043 | 1000 | 0.003 | 0.002 | 0.002 | 0.001 | 0.103 | | 0.061 | 0.073 | 0.034 | 0.709 | | 0.262 | 0.482 | 0.223 |
| DLM | 200 | 0.144 | 0.324 | 0.345 | 0.102 | 0.253 | | 0.563 | 0.604 | 0.18 | 0.376 | | 0.830 | 0.897 | 0.266 |
| EMB | 1200 | 0.457 | 0.664 | 0.610 | 0.508 | 1.25 | | 1.82 | 1.67 | 1.38 | 4.10 | | 6.02 | 5.52 | 4.48 |
| INH | 300 | 0.094 | 0.113 | 0.077 | 0.010 | 0.646 | | 0.38 | 0.339 | 0.467 | 3.87 | | 0.739 | 0.756 | 1.08 |
| LZD | 1200 | 3.32 | 3.34 | 3.60 | 4.47 | 11.8 | | 8.92 | 7.79 | 9.83 | 22.4 | | 14.2 | 11.2 | 14.2 |
| MXF | 400 | 0.304 | 1.57 | 0.508 | 0.578 | 1.0 | | 4.17 | 1.54 | 1.50 | 2.25 | | 7.33 | 3.13 | 2.66 |
| PMD | 200 | 0.838 | 1.36 | 1.34 | 0.866 | 1.23 | | 1.96 | 1.95 | 1.26 | 1.60 | | 2.55 | 2.54 | 1.65 |
| PZA | 1500 | 3.56 | 2.40 | 2.21 | 2.14 | 14.2 | | 9.0 | 8.67 | 8.08 | 30.6 | | 18.8 | 18.8 | 17.0 |
| RIF | 1200 | 0.016 | 0.012 | 0.007 | 0.004 | 4.96 | | 3.67 | 2.0 | 1.15 | 22.83 | | 16.8 | 9.28 | 5.3 |
| RPT | 1200 | 14.4 | 11.9 | 14.5 | 8.55 | 22.3 | | 17.4 | 21.9 | 10.3 | 30.0 | | 22.4 | 28.8 | 11.5 |
| SZD | 1600 | 0.126 | 0.131 | 0.097 | 0.080 | 0.646 | | 0.265 | 0.290 | 0.336 | 1.92 | | 0.381 | 0.510 | 0.768 |
| TBAJ-587 | 100 | 0.354 | 3.17 | 3.92 | 0.829 | 0.397 | | 3.56 | 4.40 | 0.838 | 0.447 | | 4.0 | 4.94 | 0.841 |
| TBAJ-876 | 100 | 0.098 | 1.12 | 1.45 | 0.216 | 0.138 | | 1.37 | 1.63 | 0.226 | 0.200 | | 1.58 | 1.77 | 0.233 |
| TBI-223 | 2400 | 0.244 | 0.166 | 0.178 | 0.208 | 4.08 | | 2.71 | 2.91 | 3.41 | 9.51 | | 6.29 | 6.76 | 7.91 |

All values represent total concentrations. Steady-state is defined by 4 months (112 days) of dosing.

*Abbreviations:* Cmax: maximum concentration, Cmin: minimum concentration, Css,avg: average concentration at steady-state.

**Table S3. Clinical site-of-disease PK metrics used for simulations**^S1^

| **Drug** | **Plasma-to-tissue partition coefficient** | | | **Plasma-to-tissue delay rate** (1/h) | | |
| --- | --- | --- | --- | --- | --- | --- |
|  | **Uninvolved lung** | **Cellular lesion** | **Caseum** | **Uninvolved lung** | **Cellular lesion** | **Caseum** |
| BDQ | 11.27 | 8.47 | 1.67 | 10 | 0.141 | 10 |
| BTZ-043 | 0.59 | 0.71 | 0.33 | 0.62 | 10.8 | 10 |
| DLM | 2.22 | 2.39 | 0.71 | 2.63 | 10 | 10 |
| EMB | 1.45 | 1.33 | 1.1 | 8.1 | 7.52 | 2.44 |
| INH | 0.586 | 0.522 | 0.719 | 0.146 | 0.19 | 0.202 |
| LZD | 0.755 | 0.659 | 0.833 | 0.409 | 0.243 | 0.252 |
| MXF | 4.14 | 1.54 | 1.51 | 0.363 | 0.77 | 0.354 |
| PMD | 1.6 | 1.59 | 1.03 | 2.44 | 10 | 10 |
| PZA | 0.635 | 0.613 | 0.571 | 1.83 | 10 | 2.12 |
| RIF | 0.737 | 0.406 | 0.232 | 10 | 10 | 10 |
| RPT | 0.78 | 0.98 | 0.46 | 0.53 | 0.82 | 0.13 |
| SZD | 0.41 | 0.45 | 0.52 | 0.132 | 0.238 | 0.533 |
| TBAJ-587 | 8.97 | 11.07 | 2.14 | 10.37 | 13.16 | 0.016 |
| TBAJ-876 | 9.92 | 11.88 | 1.65 | 0.15 | 0.08 | 0.03 |
| TBI-223 | 0.66 | 0.71 | 0.83 | 10 | 10 | 1.72 |

**Table S4. Early potency metrics used for PK-PD coverage analysis**

| **Drug** | ***In vitro***^S1^ | | | | ***In vivo*** | | | |
| --- | --- | --- | --- | --- | --- | --- | --- | --- |
|  | **MIC** (mg/L) | **macIC_90_** (mg/L) | **casMBC_50_** (mg/L) | **casMBC_90_** (mg/L) | **EC_50_** (mg/L)* | **EC_90_**  (mg/L)**^†^** | **Infection model** | **Reference** |
| BDQ | 0.06 | 0.05 | 1.17 | 2.61 | 0.052 | 0.468 | Subacute | In-house |
| BTZ-043 | 0.001 | 0.0016 | 220.9 | >220.9 | 0.005135 | 0.0463 | Chronic | ^S22^ |
| DLM | 0.002 | 0.01 | 1.07 | 4.28 | 1.02 | 9.18 | Subacute | ^S23^ |
| EMB | 0.6 | 4.09 | 104.4 | >104.4 | 0.150 | 1.35 | Acute | ^S23^ |
| INH | 0.06 | 0.04 | 0.56 | 70.22 | 0.00404 | 0.0364 | Subacute | ^S23^ |
| LZD | 0.5 | 6.75 | 172.72 | >172.72 | 9.85 | 88.65 | Subacute | In-house |
| MXF | 0.12 | 1 | 0.05 | 0.24 | 0.0000586 | 0.000527 | Subacute | ^S23^ |
| PMD | 0.04 | 1.4 | 1.08 | 25.15 | 3.46 | 1213 | Subacute | ^S23^ |
| PZA | 12.5 | 2.4 | 98.5 | 1009.09 | 7.85^‡^ | 70.65 | Acute | ^S24^ |
| RIF | 0.02 | 0.25 | 2.24 | 8.23 | 1.92 | 9.44 | Subacute | ^S23^ |
| RPT | 0.01 | 0.02 | 0.79 | 8.77 | 6.02 | 15.27 | Chronic | ^S23^ |
| SZD | 0.1 | 0.001 | 0.49 | 5.65 | 0.269 | 2.42 | Chronic | In-house |
| TBAJ-587 | 0.004 | 0.01 | 0.17 | 0.86 | 0.0362 | 0.326 | Subacute | In-house |
| TBAJ-876 | 0.001 | 0.01 | 0.03 | 0.13 | 0.0436 | 0.392 | Subacute | In-house |
| TBI-223 | 2 | 4.2 | 46.77 | >46.77 | 2.86 | 97.29 | Acute | ^S20^ |

MICs were assessed in standard media (7H9 broth, or 7H10, 7H11, or 7H12 medium), with or without enrichment.

*Estimated in mouse. Values were corrected by plasma protein binding differences between species for translation to clinical potency.

**^†^**Calculated from EC_50_ using (90/(100-90))^(1/Hill)^*EC_50_

^‡^EAUC_50_ (mg*h/L)

*Abbreviations:* casMBC_90_: concentration that kills 90% of non-replicating persisters in *ex vivo* rabbit caseum, EC_50_: *in vivo* half-maximal effective concentration, macIC_90_: concentration at which 90% of bacterial growth is inhibited in the macrophage, MIC: minimum inhibitory concentration.

**Table S5. Clinical trials and/or modeling analyses revealing efficacious doses**

| **Drug** | **Drug class** | **Efficacious daily dose(s) (mg)*** | **Current evidence** | **Reference** |
| --- | --- | --- | --- | --- |
| BDQ | Diarylquinoline | 200 | Phase 3 (Nix; NCT02333799)  Phase 3 (ZeNix; NCT03086486) | ^S25,26^ |
| BTZ-043 | DprE1 inhibitor | 1000 | Phase 1b/2a (PanACEA-BTZ-043-02; NCT04044001 | ^S27^ |
|  |  | 250, 500, 1000 | Phase 2b (DECISION; NCT05926466) | ^-^ |
| DLM^+^ | Nitroimidazole | 200 | Phase 2b (PanACEA-DECODE-01; NCT04550832)  Phase 2b (PanACEA-SUDOCU-01; NCT03959566) | ^S28,29^ |
|  |  | 400 | Phase 2 (NCT00685360) | ^S30^ |
| EMB | Other | 800, 1200, 1600 | Phase 3 (Study 31/A5349; NCT02410772) | ^S31^ |
| INH | Other | 300 | Phase 3 (Study 31/A5349; NCT02410772) | ^S31^ |
| LZD | Oxazolidinone | 600, 1200 | Phase 3 (Nix; NCT02333799)  Phase 3 (ZeNix; NCT03086486) | ^S25,26^ |
| MXF | Fluoroquinolone | 400 | Phase 3 (Study 31/A5349; NCT02410772) | ^S31^ |
| PMD | Nitroimidazole | 100, 200 | Phase 2b (NC-002; NCT01498419) | ^S32^ |
|  |  | 200 | Phase 3 (Nix; NCT02333799)  Phase 3 (ZeNix; NCT03086486) | ^S25,26^ |
| PZA | Other | 1000, 1500, 2000 | Phase 3 (Study 31/A5349; NCT02410772) | ^S31^ |
|  |  | 1000 | Modeling | ^S16^ |
| RIF | Rifamycin | 600 | Phase 3 (Study 31/A5349; NCT02410772) | ^S31^ |
|  |  | 600 | Modeling | ^S33^ |
|  |  | 600, 900, 1200 | Phase 2b (HIRIF; NCT01408914) | ^S34,35^ |
| RPT | Rifamycin | 1200 | Phase 2b (Study 29X; NCT00694629)  Phase 3 (Study 31/A5349; NCT02410772) | ^S31,36^ |
| SZD | Oxazolidinone | 600, 1200, 1600 | Phase 2a+ (RAD-TB/A5409; NCT06192160)  Phase 2b (PanACEA-SUDOCU-01; NCT03959566) | ^S29,37^ |
| TBAJ-587 | Diarylquinoline | 100 | Phase 1 (NCT04890535); Modeling | ^S38^ |
| TBAJ-876 | Diarylquinoline | 25, 50, 100 | Phase 2b/c (NC-009; NCT06058299) | ^-^ |
| TBI-223 | Oxazolidinone | 1200, 2400 | Modeling | ^S20^ |

*mg/kg doses were converted to flat doses using a body weight of 60 kg.

^+^Twice-daily doses were converted to once-daily doses.

**Table S6. Clinical caseum css,avg vs. casMBC_90_**

| **Drug** | **Dose (mg)** | **Css,avg/casMBC_90_** |
| --- | --- | --- |
| BDQ | 200 | **1.22** |
| BTZ-043 | 1000 | 0.00015 |
| DLM | 200 | 0.042 |
| EMB | 1200 | 0.013 |
| INH | 300 | 0.007 |
| LZD | 1200 | 0.057 |
| MXF | 400 | **6.27** |
| PMD | 200 | 0.050 |
| PZA | 1500 | 0.008 |
| RIF | 1200 | 0.140 |
| RPT | 1200 | **1.17** |
| SZD | 1600 | 0.059 |
| TBAJ-587 | 100 | **0.97** |
| TBAJ-876 | 50 | **1.74** |
| TBI-223 | 2400 | 0.073 |

**Table S7. Mouse PK data and models used for simulations**

| **Drug** | **Simulated model structure** | **Model reference** | **fu** |
| --- | --- | --- | --- |
| BDQ | 3 compartment disposition; first-order absorption; first-order elimination | In-house | 0.000047 |
| BTZ-043 | 2 compartment disposition; Michaelis-Menten absorption; first-order elimination | ^S22^ | 0.029 |
| DLM | 1 compartment disposition; first-order absorption; first-order elimination | ^S23^ | 0.005^S7^ |
| EMB | 2 compartment disposition; first-order absorption; first-order elimination | ^S23^ | 0.879^S23^ |
| INH | 2 compartment disposition; first-order absorption; Michaelis-Menten elimination | ^S23^ | 0.58^S39^ |
| LZD | 2 compartment disposition; first-order absorption; Michaelis-Menten elimination | ^S23^ | 0.7^S40^ |
| MXF | 2 compartment disposition; first-order absorption; first-order elimination | ^S23^ | 0.69^S13^ |
| PMD | 2 compartment disposition; first-order absorption; dose-dependent F; Michaelis-Menten elimination | ^S23^ | 0.09^S41^ |
| PZA | 2 compartment disposition; first-order absorption; dose-dependent F; Michaelis-Menten elimination | ^S23^ | 1 |
| RIF | 2 compartment disposition; first-order absorption; dose-dependent F; Michaelis-Menten elimination | ^S23^ | 0.032^S42^ |
| RPT | 1 compartment disposition; first-order absorption; Michaelis-Menten elimination | ^S23^ | 0.009 |
| SZD | 1 compartment disposition; first-order absorption; first-order elimination | In-house | 0.458 |
| TBAJ-587 | 3 compartment disposition; first-order absorption; first-order elimination dose-dependent F | In-house | 0.00024 |
| TBAJ-876 | 3 compartment disposition; first-order absorption; first-order elimination | In-house | 0.00023 |
| TBI-223 | 2 compartment disposition; first-order absorption; Michaelis-Menten elimination; allometric scaling by WT | ^S20^ | 0.671^S20^ |

*Abbreviations:* F: bioavailability, fu: fraction unbound in plasma, WT: body weight.

**Table S8. Mouse vs. human plasma PK**

| **Drug** | **Mouse** | | **Human** | |
| --- | --- | --- | --- | --- |
|  | **Dose (mg/kg)** | **AUCss,_0-24_ (mg*h/L)** | **Dose (mg)** | **AUCss,_0-24_ (mg*h/L)** |
| BDQ | 75 | 53.4 | 200 | 45.7 |
| BTZ-043 | 15 | 2.52 | 1000 | 2.48 |
| DLM | 4 | 6.59 | 200 | 6.08 |
| EMB | 100 | 25 | 1200 | 30.1 |
| INH | 15 | 13.3 | 300 | 15.5 |
| LZD | 125 | 258 | 1200 | 283 |
| MXF | 150 | 25.2 | 400 | 24 |
| PMD | 25 | 33.3 | 200 | 29.5 |
| PZA | 300 | 366 | 1500 | 340 |
| RIF | 10 | 119 | 1200 | 119 |
| RPT | 10 | 610 | 1200 | 536 |
| SZD | 200 | 16.8 | 1600 | 15.5 |
| TBAJ-587 | 12.5 | 8.84 | 100 | 9.53 |
| TBAJ-876 | 5 | 3.31 | 50 | 3.3 |
| TBI-223 | 400 | 83.2 | 2400 | 98 |

Steady-state is defined by 4 months (112 days) of dosing.

**Table S9. Mouse plasma css,avg vs. EC_50_**

| **Drug** | **Dose (mg/kg)** | **Css,avg/EC_50_** |
| --- | --- | --- |
| BDQ | 75 | **42.8** |
| BTZ-043 | 15 | **20.5** |
| DLM | 4 | 0.27 |
| EMB | 100 | **6.94** |
| INH | 15 | **137.2** |
| LZD | 125 | 1.09 |
| MXF | 150 | **17918** |
| PMD | 25 | 0.4 |
| PZA | 300 | 1.94 |
| RIF | 10 | 2.58 |
| RPT | 10 | **3.16** |
| SZD | 200 | 2.6 |
| TBAJ-587 | 12.5 | **10.2** |
| TBAJ-876 | 5 | **3.16** |
| TBI-223 | 400 | 1.21 |

**Table S10. Comparison of predicted effective dose range vs. current efficacious dose(s) across all PK-PD targets**

| **Drug** | **Clinical efficacious daily dose (mg)** | **Predicted ED_50_-ED_90_** | | | | |
| --- | --- | --- | --- | --- | --- | --- |
|  |  | **Total plasma PK over MIC** | **Unbound plasma PK over MIC** | **Caseum PK over casMBC_90_** | **Cellular lesion PK over EC_90_** | **Cellular lesion PK over EC_50_** |
| BDQ | 200 | 8-10 | >10000 | 153-196 | 8-10 | <1 |
| BTZ-043 | 250-1750 | 55-232 | 992-4190 | >10000 | 1850-7700 | 208-850 |
| DLM | 200 | 3-5 | 330-504 | 4920-7533 | 3129-4800 | 352-537 |
| EMB | 800-1600 | 1041-1474 | 1189-1681 | >10000 | 1756-2485 | 193-279 |
| INH | 300 | 114-183 | 148-274 | >10000 | 38-84 | 4-9 |
| LZD | 600-1200 | 136-354 | 185-464 | 2850-2890 | 2726-2785 | 1371-1727 |
| MXF | 400 | 65-167 | 117-261 | 74-170 | <1 | <1 |
| PMD | 200 | 3-5 | 32-50 | 9575->10000 | >10000 | 244-361 |
| PZA | 1000-2000 | 1271-4750 | 1293-4845 | >10000 | >10000 | 1313-4900 |
| RIF | 600-1800 | 30-750 | 334-2963 | 5800-9500 | 3000-7150 | 1333-5250 |
| RPT | 1200-1800 | <1 | 183-275 | 1009-1211 | 844-1200 | 336-465 |
| SZD | 800-1600 | 605-1208 | 1054-1890 | 5448-5573 | 4704-5023 | 1555-2464 |
| TBAJ-587 | 25-400 | <1 | 5417-6449 | 150-190 | 8-10 | <1 |
| TBAJ-876 | 25-100 | <1 | 919-1157 | 30-46 | 30-46 | 3-5 |
| TBI-223 | 1200-2400 | 1675-5067 | 2350-5997 | >10000 | >10000 | 2867-6550 |

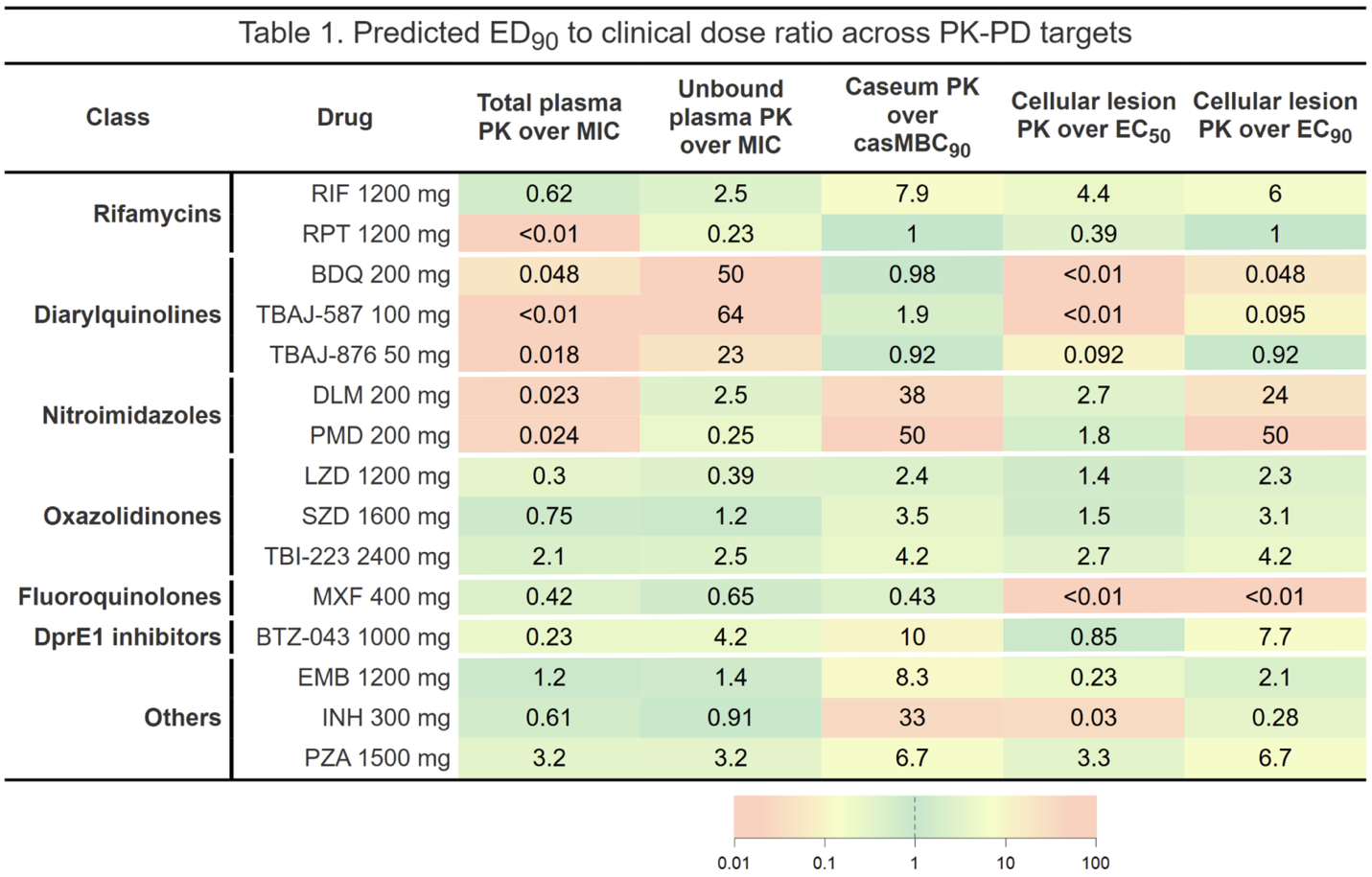

**Figure S1. Ratio of predicted ED_90_ over clinical efficacious dose across all PK-PD targets.** Ratios were log_10_-transformed and mapped to a symmetrical green-to-red color scale. Values close to 1 are shown in green, indicating minimal deviation. Large deviations in either direction are shaded in red.
